# 20+ years of mass drug administration against urogenital schistosomiasis on the Zanzibar islands: Is praziquantel still efficacious?

**DOI:** 10.64898/2026.08.13.26360376

**Authors:** Stefanie Knopp, Norbert J. van Dijk, Naomi C. Ndum, Tom Pennance, Mohammed N. Ali, Khamis R. Suleiman, Matthew Denwood, Saleh Juma, Shaali M. Ame, Aidan M. Emery, Bonnie L. Webster, Luc E. Coffeng, Said M. Ali

## Abstract

**Background:** On the Zanzibar islands, Tanzania, mass drug administration (MDA) with praziquantel against *Schistosoma haematobium* infections has been implemented regularly since the early 2000s. Elimination of schistosomiasis as a public health problem has been achieved in most areas, with the goal of interruption of transmission. The RESIST project investigates the extent to which MDA-driven selection for praziquantel resistance is occurring and contributing to persistent transmission hotspots.

**Methodology:** As part of RESIST, the efficacy of praziquantel treatment was assessed in two schools on Pemba between October 2024 and March 2025. Longitudinal parasitological surveys were conducted before and two weeks after school-based MDA with praziquantel (40 mg/kg). Up to six urine samples per study participant were collected on different days pre- and post-MDA and examined for *S. haematobium* eggs by urine filtration microscopy. In a per-protocol analysis, drug efficacy was categorised as adequate, inconclusive, or reduced, based on hypothesis testing. A generalized linear mixed model was used to assess the association between pre-MDA infection intensity and drug efficacy at the individual level.

**Principal findings:** Pre-MDA, *S. haematobium* prevalence was 14.3% (62/434) in School 1 and 37.8% (233/617) in School 2. Post-MDA prevalence was 0.5% (2/434) and 9.6% (59/617), respectively. Egg reduction rates were 99.8% (90% confidence interval (CI): 99.2-100%) in School 1 and 94.8% (90% CI: 88.6-98.6%) in School 2, which were classified as adequate and inconclusive, respectively. In School 2, drug efficacy at the individual level was negatively associated with pre-MDA infection intensity.

**Conclusion/significance:** The efficacy of praziquantel on Pemba remains higher than the 90% threshold for optimal drug efficacy set by the World Health Organization. While the inconclusive efficacy results for School 2 can at least partially be explained by the variation in the participants’ pre-MDA infection intensities, further investigations are warranted to account for the disparity in praziquantel efficacy on Pemba.

**Trial registration:** ISRCTN, ISRCTN59331501. Registered 24 October 2024, https://www.isrctn.com/ISRCTN59331501.

**Author Summary:** On the Zanzibar islands, Tanzania, mass drug administration (MDA) with praziquantel against schistosomiasis has been implemented regularly since the early 2000s. Each year, several hundred thousand people are treated. Consequently, in most areas, schistosomiasis has been successfully reduced, but some hotspots of transmission persist. The RESIST project investigates if MDA-driven selection of praziquantel resistance is occurring and contributing to the existence of hotspots. We assessed praziquantel efficacy in two schools on Pemba. Parasitological surveys were conducted before and two weeks after school-based MDA. In each survey, up to six urine samples per participant were collected and examined for *Schistosoma haematobium* eggs by microscopy. Results showed that praziquantel efficacy in both schools was higher than 90%. According to the current World Health Organization’s manual for evaluating drug efficacy against parasitic worm infections, this means that praziquantel remains efficacious. Since the recommended evaluation method in this manual is statistically flawed, we applied formal hypothesis testing, demonstrating that praziquantel efficacy was adequate in School 1 but inconclusive in School 2. The lower efficacy in School 2 can at least partially be explained by individuals’ high intensity infections pre-MDA, but further investigations are warranted to account for the disparity in praziquantel efficacy on Pemba.

## Introduction

Schistosomiasis is a neglected tropical disease (NTD). It is caused by infection with parasitic blood flukes of the genus *Schistosoma* with transmission reported in 79 countries worldwide [1,2]. Disease in humans is mainly caused by three species: *S. haematobium* infection can result in urogenital schistosomiasis, while *S. mansoni* and *S. japonicum* infection can result in intestinal schistosomiasis [2,3]. The parasite transmission cycle includes infected human hosts excreting parasite eggs in urine (*S. haematobium*) or feces (*S. mansoni* and *S. japonicum*) into freshwater, intermediate freshwater host snails wherein the parasites replicate asexually, and parasite larvae released by the snails, which can penetrate the skin of human hosts that are in contact with freshwater and develop into reproducing *Schistosoma* adult worms within the human body [3,4]. Chronic *Schistosoma* infection can lead to considerable morbidity [3,4]. The global burden of disease attributable to schistosomiasis was estimated at 1.75 million disability-adjusted life years in 2021 [5]. To control schistosomiasis and prevent morbidity, the World Health Organization (WHO) recommends the periodic administration of praziquantel (40 mg/kg), to populations at risk, via preventive chemotherapy programs, alongside other interventions [1,2].

Praziquantel has been the drug of choice for treating schistosomiasis for almost 50 years [6,7]. It has been delivered via preventive chemotherapy programs since the late 1980s/early 1990s [8,9], and was recommended by the WHO for large-scale mass drug administration (MDA) in 2006 [10]. Merck began praziquantel donations, in coordination with the WHO, in 2007 [11]. Since then, preventive chemotherapy programs distributing praziquantel by MDA have been scaled up significantly, reducing schistosomiasis prevalence levels in many endemic countries [12].

However, the increased use of praziquantel in preventive chemotherapy programs also elevated the risk of *Schistosoma* parasites developing drug resistance. In response, the WHO published revised guidelines for monitoring drug efficacy against schistosomiasis in 2013 [13]. Egg reduction rates (ERR), referring to the reduction of the number of *Schistosoma* eggs in urine or feces from before to after treatment, were considered the best measure for drug efficacy [14], with a threshold of point-estimate ≥90% ERR suggested for sufficient efficacy [13]. Unfortunately, detecting drug resistance based on ERR is not straightforward as several factors can confound drug efficacy (e.g. food intake, food types, drug interactions and comorbidities) and alter daily egg excretion (e.g. host biological rhythms, fluid intake, or physical exercises) [15,16]. As such, to date there is no published and unequivocal evidence of schistosomes developing resistance to praziquantel in endemic areas where praziquantel has been provided for multiple years [7]. However, reduced efficacy of praziquantel based on cure rates and ERR has been reported for *S. mansoni* in several East African countries [17,18], and several studies on laboratory strains of *S. mansoni* have identified mutations in the praziquantel target (transient receptor potential ion channel *Sm*TRPM_PZQ_) that can cause a complete lack of response of the channel to praziquantel [19–25]. So far, most research on praziquantel efficacy and resistance has focused on *S. mansoni*, whereas few field-based and no laboratory-based investigations have been done on *S. haematobium*.

On the Zanzibar islands, United Republic of Tanzania, the only endemic *Schistosoma* species infecting humans is *S. haematobium* [26–28]. The islands are among the few endemic areas where pilot preventive chemotherapy programs to control schistosomiasis commenced as early as the late 1980s, with large-scale regular MDAs in schools introduced in the early 2000s [9,28]. Efforts to eliminate schistosomiasis from the islands started in 2012 [29]. These efforts included biannual MDAs in schools and communities, in combination with snail control or behavior change interventions, and were implemented from 2012 to 2016 as part of the Zanzibar Elimination of Schistosomiasis Transmission (ZEST) project [29–31]. From 2017 to 2024, with a gap in 2019, MDA was administered at least annually. Since the onset of large-scale interventions against schistosomiasis on the Zanzibar islands in the 1980s, the prevalence of *S. haematobium* has declined to low levels [28], with only 3.4% of schoolchildren and 0.4% of adults infected across 90 communities in 2020 [32]. However, while to date many areas on the islands have zero or very low prevalence levels, some areas with a prevalence of at least 10% remain and are considered hotspots of transmission [32]. Rurality, proximity of houses to water bodies containing intermediate host snails of *S. haematobium*, and a low economic standard of households clustered in certain areas partially explain the existence of hotspots [33,34]. However, the intense drug pressure imposed by more than 20 years of praziquantel MDAs on the Zanzibar islands presents uncertainties regarding praziquantel efficacy and possible resistance in these hotspots. As part of the RESIST project [35], this study assessed praziquantel efficacy in two hotspot schools on Pemba Island, Zanzibar. We assessed the ERR in each school using parasitological methods, applied a statistical framework to classify drug efficacy, and analysed ERR variation between students using a generalized linear mixed model.

## Methods

### Ethics

The RESIST study protocol received ethical approval from the ethics committee of Northwestern and Central Switzerland (Ethikkommission Nordwest-und Zentralschweiz; EKNZ) on August 13, 2024 (AO_2024_00084) and from the Zanzibar Health Research Institute (ZAHRI) on September 24, 2024 (ZAHREC/01/PR/Sept/2024/33). The study was prospectively registered at ISRCTN (ISRCTN59331501), and the study protocol has been published [35].

Before the fieldwork started, meetings were held with delegates from the Zanzibar Ministry of Health, district health authorities, community leaders, school principals and parents to explain the purpose and procedures of the study and to discuss questions and concerns about the study. The study was also explained in detail to the students of the two schools that were eligible for participation. In addition, all eligible students received an information sheet in Kiswahili (local language) about the study procedure, including the telephone number of the local principal investigator and secretary of ZAHRI, respectively, whom parents could call if they had any questions. Students could only participate if they submitted a written informed consent form signed by their parent/legal guardian. Additionally, if students were 12 years or older, they were requested to sign a written assent form for their own participation.

### Study site

The RESIST drug efficacy study was conducted in two public primary schools on Pemba, the northernmost island of the semi-autonomous Zanzibar archipelago. Pemba is divided into two regions (North and South) and four districts (Chake, Micheweni, Mkoani and Wete). The population of Pemba was estimated at 543,441 inhabitants at the last census conducted in 2022 [36]. The net enrolment rate for primary schools in the South region was 96.7% in 2022 [36]. Our two study schools were located in Chake district (South region) and were known as historical *S. haematobium* hotspots from earlier ZEST surveys, where they had a prevalence >10% before the ZEST interventions started in 2012 and persisted at levels >5% throughout the five years of the ZEST project, until 2017 [30]. Based on more recent surveys conducted by the NTD program of the Zanzibar Ministry of Health, we expected the prevalence of *S. haematobium* infection in these schools to be at least 5%. Before our study was implemented in late 2024 and early 2025, the last MDA in the two schools had been conducted in May 2023.

### Study design and sample size determination

The RESIST drug efficacy study was designed as a longitudinal study of repeated parasitological surveys of students attending primary school [35]. We aimed to collect quintuple urine samples (1/day) over five different days from each participating student in the two study schools, both before and two weeks after MDA with praziquantel, to assess praziquantel drug efficacy.

The approach for sample size calculation has been described in detail in the published study protocol [35]. Briefly, the study was powered for non-inferiority testing of praziquantel efficacy (which results in a sample size that is automatically sufficient for inferiority testing as well), assuming an expected efficacy of 98% and a non-inferiority margin of five percentage points. We set an alpha of 0.10 for both the non-inferiority and inferiority test (producing an overall risk of a type I error of 0.05) and a desired power of 90% (risk of type II error of 0.10). Considering an expected pre-MDA prevalence of egg-positivity of 5%, and the typical high overdispersion of egg counts between and within individuals (upon repeated testing over multiple days), we determined that quintuple testing of 500 individuals (over five days) per school, both before and after praziquantel treatment, would meet the desired maximum type I and II errors.

To reach a total of 500 students per school for final inclusion in the efficacy study and accounting for a daily drop-out rate of 10% during sample collection, we aimed to enrol 800 students in each school.

### Participant recruitment and sample collection

Recruitment for the drug efficacy study in School 1 started on 04/11/2024 and ended on 29/11/2024. Recruitment in School 2 started on 31/01/2025 and ended on 18/03/2026. Students were eligible to participate in the study when they i) attended school grades 1-7, ii) were aged 5-16 years, iii) submitted written informed consent signed by their parent/legal guardian, and iv) submitted written assent signed by themselves if 12-16 years old.

The longitudinal study of repeated parasitological surveys of students before and after praziquantel MDA was implemented as follows. Three to one weeks before MDA, on Day 0, all present students from grade 1-7 were registered and received a personal identifier code. Since the number of attending students in each school was smaller than 800, all students were eligible to participate. On Day 1, following submission of the relevant signed consent/assent forms, students received a transparent, screw-cap plastic container (100 ml) labelled with their personal identifier code and were asked to collect their own urine between 10.00 am and 2.00 pm. The urine collection procedure was repeated on Days 2-5, so that each student provided a total of five urine samples (1/day) before MDA. Next, MDA with praziquantel was conducted as described below. Two weeks after MDA, the sampling procedures from Days 1-5 were repeated. To reduce dropout and reach the needed sample size, we conducted supplementary sample collection days before and after MDA, and one supplementary day of treatment for the study participants who were absent on the day scheduled for MDA. This treatment “mop-up” was conducted on the day after the scheduled MDA.

### Mass drug administration

The MDA, which included treatment of students with praziquantel against schistosomiasis and with albendazole against soil-transmitted helminthiases, was conducted by staff of the RESIST study team and the Pemba NTD Unit of the Zanzibar Ministry of Health, in collaboration with the schoolteachers.

Around 7.00 am, all present students received a cup of porridge to enhance the bioavailability of the anthelminthics provided [37], and then gathered in their classrooms to eat. There, each present student received one albendazole tablet (400 mg; GlaxoSmithKline) to chew and swallow. Subsequently, the students registered for the RESIST study were called one by one by name, measured with a paper praziquantel dose pole, and given the number of praziquantel tablets (600 mg; Merck) required according to their height [10,38,39]. Depending on the required dose and to ease the swallowing of the large tablets, some students had tablets broken in half as part of their dose. Together with the praziquantel tablets, each student received a cup of drinking water, and it was directly observed if the student swallowed all provided praziquantel tablets completely. The number of praziquantel tablets each student received and swallowed was recorded, as well as whether the student vomited during tablet intake or within one hour after treatment. Additional students that were not registered for the RESIST study but present on the day of MDA received praziquantel after the RESIST study participants.

### Urine filtration and *S. haematobium* egg microscopy

Urine samples were transferred from the school to the Public Health Laboratory-Ivo de Carneri (PHL-IdC) within two hours after collection and processed immediately. Each urine sample was mixed, and 10 ml were filtered through a SEFAR 13 mm diameter cold punched filter disc (Sefar, United Kingdom; cat. no. G060-1021-001-00) held within a 13mm Swinnex filter holder (Merck, Germany; cat no. SX0001300) attached to a plastic syringe. Each individual filter disc was transferred to a separate microscope slide that was labelled with the participant identifier code and covered with a piece of hydrophilic cellophane soaked in glycerol. *S. haematobium* eggs were stained by adding a few drops of 10% Lugol’s iodine solution on top of the cellophane. The filter area was examined for the presence and number of *S. haematobium* eggs with a compound light microscope by experienced laboratory technicians. The presence and number of *S. haematobium* eggs per 10 ml of urine were recorded on paper case report forms, for each study participant on each day.

### Data management

Demographic data collected during registration of the students in their schools were recorded using Open Data Kit (ODK, www.opendatakit.org) installed on Samsung Galaxy Tab A 19 tablets. Treatment data collected during MDA in the schools and *S. haematobium* egg counts recorded in the laboratory were captured on paper forms and double-entered in Microsoft Excel spreadsheets (version 2016) by two experienced members of the study team. Data cleaning was conducted using StataBE19. Any data mismatches detected in the double-entry were compared with entries in the original paper forms and corrected. For statistical analyses, coded registration, treatment and urine microscopy data were merged by individual participant identifier codes.

### Statistical analysis

Data from School 1 and 2 were analyzed separately using R version 4.5.1 [40]. As this was a drug efficacy monitoring study, we used a per-protocol (PP) analysis where only participants who provided at least one urine sample both pre- and post-MDA and swallowed all provided praziquantel tablets (without vomiting) during MDA were included. To ensure that the PP analysis outcomes would not be affected by potential attrition bias, we compared the age, sex and pre-MDA infection status distributions of the PP cohort with those of the dropped-out students. Age and sex were compared using the standardized mean difference (SMD), where SMD≤0.2 was considered negligible to small, 0.2<SMD≤0.5 as moderate, and SMD>0.5 as large. Pre-MDA infection status was compared descriptively.

Pre- and post-MDA prevalences of *S. haematobium* infections were calculated considering students to be infected when they had at least one egg-positive urine sample pre- or post-MDA, respectively. The 95% confidence interval (CI) of the infection prevalences was calculated using the Wilson score interval. Infection intensity was categorized as light infections (1-49 eggs/10 ml urine) and heavy infections (≥50 eggs/10 ml urine) based on a student’s arithmetic mean egg count pre- and post-MDA.

The primary outcome measure of the efficacy of praziquantel in each school was the ERR, which was calculated using the arithmetic mean egg count pre- and post-MDA as recommended by the current WHO guidelines [13]: 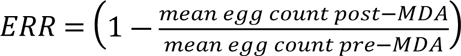 · 100%. The school-level mean egg counts were calculated as the average of the students’ arithmetic mean egg counts. We also calculated the CR for each school as a secondary measure of drug efficacy: 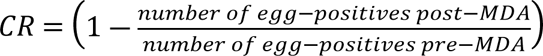· 100%. For both the ERRs and CRs, we calculated the 90% CI through a two-stage bootstrap approach: first, individuals were randomly sampled with replacement, followed by sampling with replacement of pre- and post-MDA egg counts per sampled individual. The sampled counts were then used to calculate the bootstrap ERR and CR. This procedure was repeated 1,000 times to determine the 5% and 95% percentiles of the ERR and CR sampling distributions.

To assess if the efficacy of praziquantel was either reduced or not inferior to the expected efficacy, we performed inferiority and non-inferiority testing, respectively, following the statistical framework suggested by Denwood *et al.* (S1 Fig) [41]. We assumed an expected praziquantel efficacy *T_E_* (i.e., ERR) of 98% with a non-inferiority margin (`δ`) of five percentage points. The null hypothesis of the inferiority test is that the observed praziquantel efficacy is equal to or greater than the expected praziquantel efficacy of 98%, which is rejected when the upper confidence limit of the efficacy is below 98%. For the non-inferiority test, the null hypothesis is that the observed efficacy is below the lower efficacy threshold *T_L_*= *T_E_* - `δ,` which in our study was 98% - 5% = 93%. This null hypothesis is rejected when the lower confidence limit of the observed praziquantel efficacy is equal to or above 93%. Using this statistical framework, we classified praziquantel efficacy as follows. If the null hypothesis for the non-inferiority test was rejected and the null hypothesis for the inferiority test was not rejected, praziquantel efficacy was considered “adequate”. If the null hypothesis of the inferiority test was rejected, praziquantel efficacy was considered “reduced”, even if the null hypothesis for the non-inferiority test was also rejected. If neither of the two null hypotheses was rejected, praziquantel efficacy was considered “inconclusive”. As both tests are independent and one-sided, they were performed using the bootstrap-based 90% CI of the ERR, which resulted in an overall type I error rate (alpha) of 0.05.

We also evaluated whether treatment efficacy at the individual level was associated with the intensity of infection pre-MDA. Stratifying students by observed pre-MDA egg counts and comparing bootstrap-based ERRs between strata was found not to be appropriate for this purpose, as conditioning on baseline measurements would introduce bias due to regression to the mean. Therefore, we employed a Bayesian lognormal-Poisson generalized linear mixed model (GLMM) to account for random variability in observed egg counts. We considered two sources of egg count variability, each following a log-normal distribution: i) variation in mean egg intensity between individuals, and ii) day-to-day variation in mean egg intensity within an individual. Conditional on the random effects for variation between and within individuals, egg counts were assumed to be Poisson-distributed.

We used two GLMM versions in which treatment effect was implemented in different ways. First, we modelled a common population treatment effect *β* that applied to all individuals. Treatment was included as a fixed effect in the log-linear predictor, so that exp(*β*) represented the ratio between post- *versus* pre-MDA expected geometric mean egg counts. The ERR was therefore defined as 1 ― exp(*β*). Next, we evaluated potential treatment effect modification by pre-MDA infection intensity with a second GLMM version that modelled individual-specific treatment effects as *β_i_* = ― exp(*β*_0_ + *β*_1_*u_i_*). Here, ― exp(*β*_0_) represented the ERR in an average individual, and *β*_1_ quantified the association between the individual treatment effect *β_i_* and the individual random effect *u_i_*, which was the departure of the individual expected egg count on the logarithmic scale. The individual ERR in geometric mean count was defined as 1 ― exp(*β_i_*). We used the negated exponential function for *β_i_* to only allow for negative effect values (and thus positive individual ERRs).

We fitted the GLMMs for population and individual treatment effects separately for each school, using pre-MDA data from all students (irrespective of their treatment status) and post-MDA data from students in the PP cohort. This data selection ensured maximum data availability for the model estimation of the random effects without biasing the treatment effect estimation. Posterior inference was obtained using Hamiltonian Monte Carlo implemented in Stan via the *rstan* package in R [42]. Four Markov chain Monte Carlo chains were run for 4,000 iterations each, with the first 2,000 iterations discarded as warm-up. Uncertainty was quantified in terms of central 90% Bayesian credible intervals (BCI).

## Results

### Participant’s characteristics

A total of 770 students and 748 students were registered in School 1 and 2, respectively (Fig 1). All registered students were aged 5-16 years, except for one 17-year-old student from School 1, who was considered non-eligible and therefore excluded from the analysis. At least one pre-MDA urine sample was submitted by 730 students in School 1 and by 688 students in School 2, respectively. Among them, 108 and 49 students in School 1 and School 2, respectively, did not receive praziquantel due to absence during MDA and the supplementary treatment day, and 188 and 22 additional students, respectively, did not submit any urine sample post-MDA. Hence, a total of 434 students from School 1 and 617 students from School 2 were considered for PP analyses. Of note, in the PP cohort, there were 152 students pre-MDA and nine students post-MDA, respectively, who submitted six urine samples across the five original collection days and the supplementary days. For these participants, the results of all six urine samples were considered in the analyses to avoid loss of available data. In the PP cohort, 85.3% (370/434) and 46.8% (203/434) of students in School 1 submitted at least five urine samples pre- and post-MDA, respectively. In School 2, 98.7% (609/617) and 76.5%, (472/617) of students submitted at least five urine samples pre- and post-MDA, respectively.

**Fig 1.**
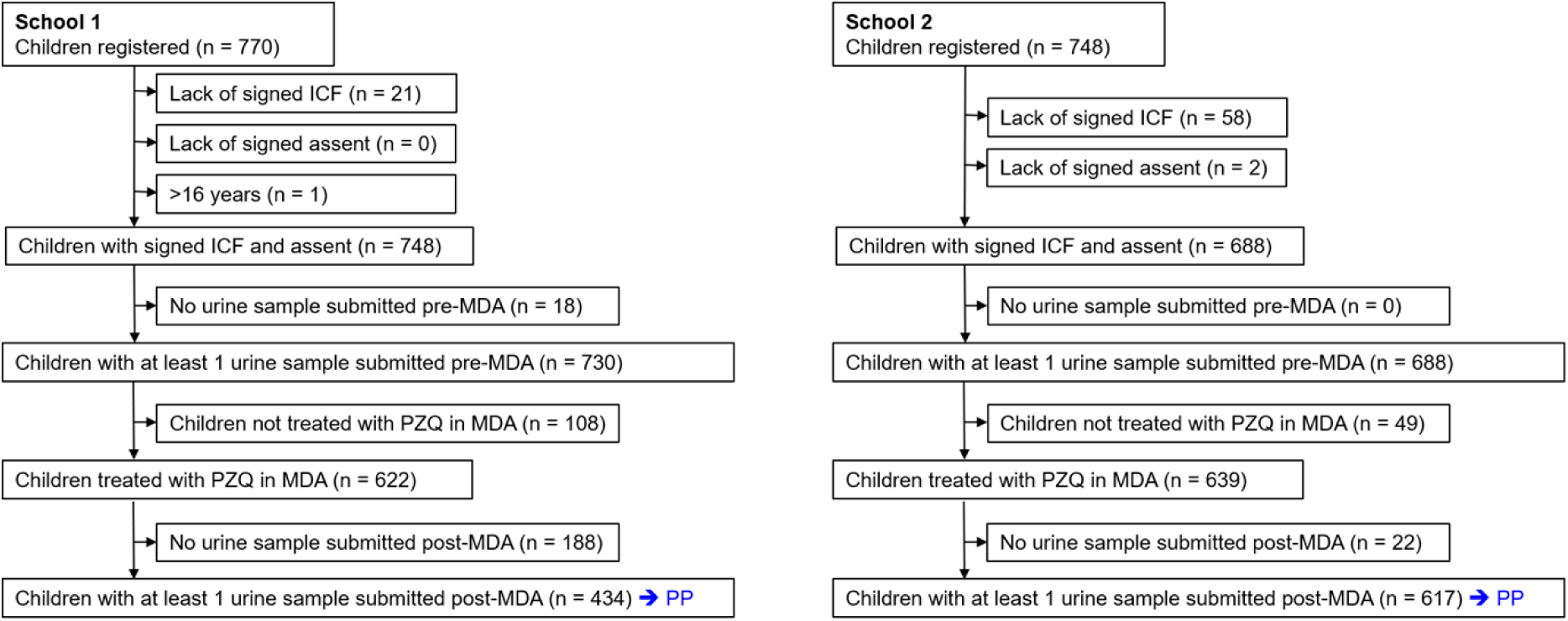
Flow-diagram of participants from two schools in the RESIST praziquantel efficacy study on Pemba, Tanzania. ICF: informed consent form; MDA: mass drug administration; PP: per protocol; PZQ: praziquantel.

Overall, there were no major differences between the baseline characteristics of the PP cohorts and drop-outs (Table 1). There was a negligible difference in age for School 1 (SMD = 0.04), while in School 2 the drop-out rate was moderately higher among younger students (SMD = 0.22). In School 1, the drop-out rate was higher among boys than girls (SMD = 0.37), whereas in School 2 the drop-out rate was equal among both sexes (SMD = 0.01). Pre-MDA infection status was comparable between the PP cohorts and drop-outs for both schools.

**Table 1.**
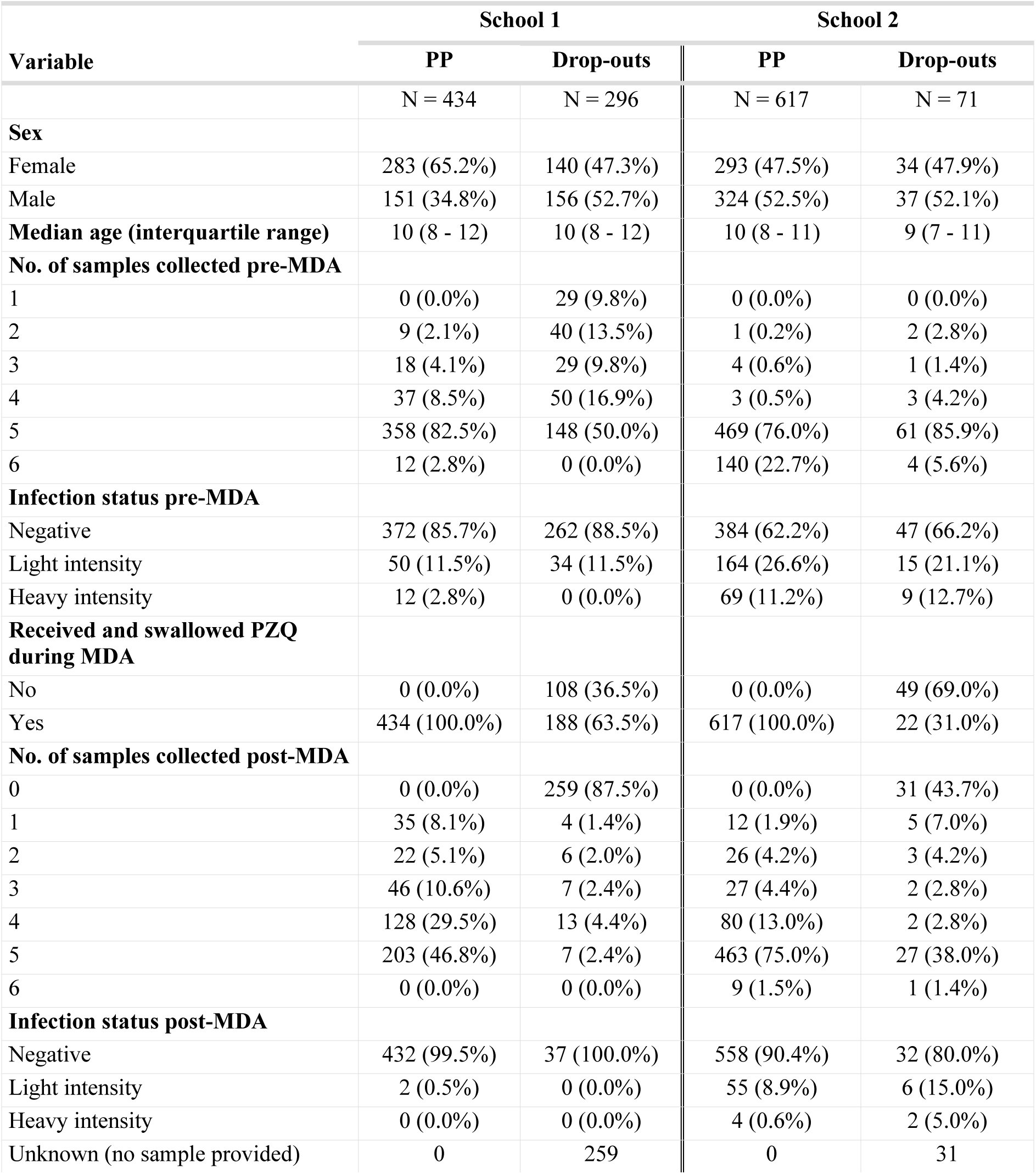
Characteristics of participants from two schools in the RESIST praziquantel efficacy study on Pemba, Tanzania. MDA: mass drug administration; PP: per protocol; PZQ: praziquantel. Light intensity infection: 1 – 49 *S. haematobium* eggs/ 10 ml urine [43], based on the arithmetic mean egg counts of all samples provided by a study participant pre-or post-MDA. Heavy intensity infection: ≥50 *S. haematobium* eggs/ 10 ml urine [43], based on the arithmetic mean egg counts of all samples provided by a study participant pre- or post-MDA.

### Change in *S. haematobium* prevalence and intensity of infection by mass drug administration in two schools

Prevalence and infection intensity pre- and post-MDA in each school, are indicated in Fig 2. In School 1, the pre-MDA *S. haematobium* prevalence was 14.3% (62/434), with 11.5% (50/434) of students having a light intensity infection and 2.8% (12/434) having a heavy intensity infection. Two weeks post-MDA, the prevalence was 0.5% (2/434) with both infections being of light intensity. In School 2, the pre-MDA *S. haematobium* prevalence was 37.8% (233/617), with 26.6% (164/617) of students having a light and 11.2% (69/617) having a heavy intensity infection. Two weeks post-MDA, the prevalence was 9.6% (59/617), with 8.9% (55/617) of the students having a light intensity and 0.6% (4/617) having a heavy intensity infection.

**Fig 2.**
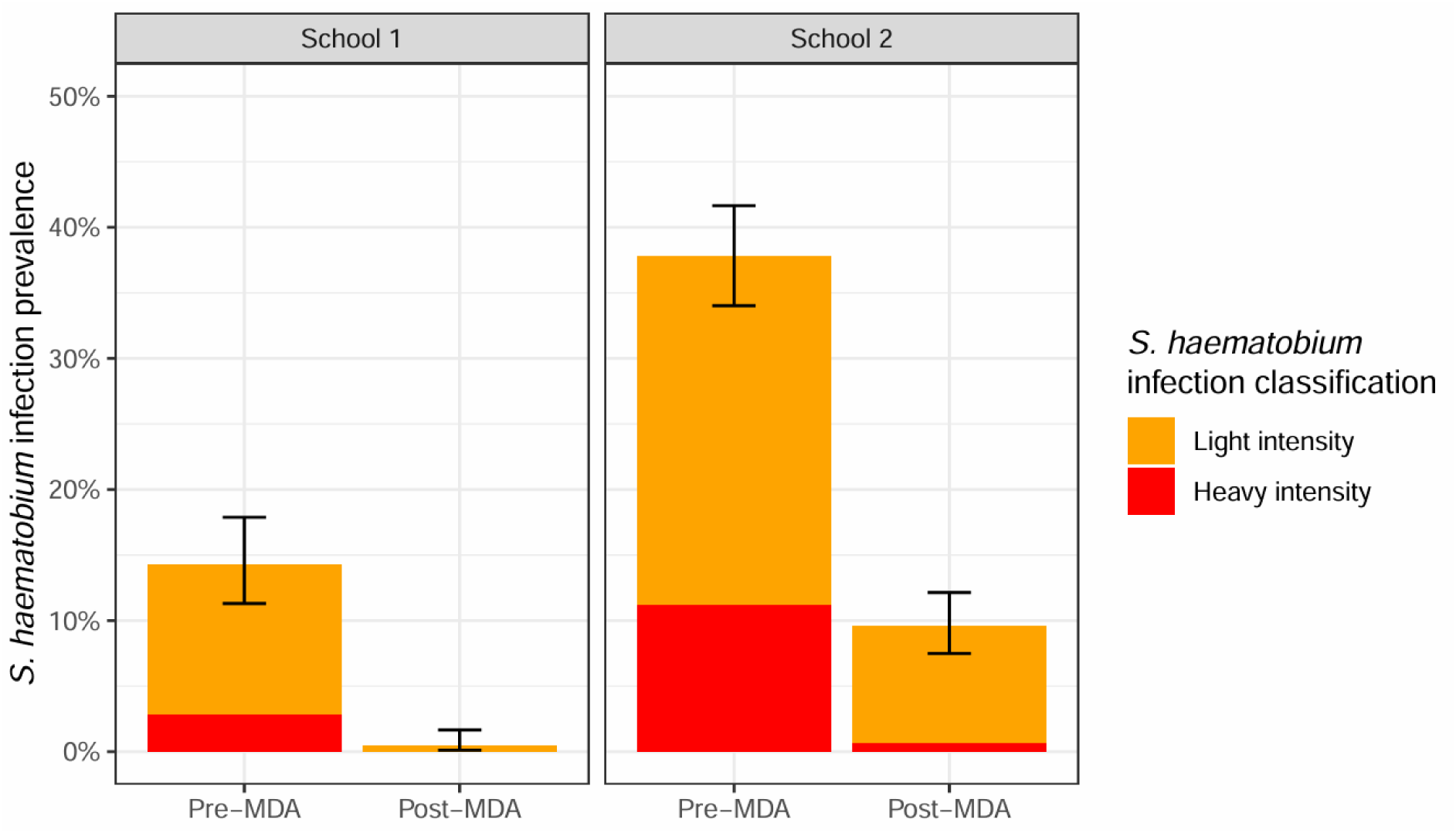
*Schistosoma haematobium* infection prevalence and intensity pre- and post-MDA in the two study schools in Pemba, Tanzania. Light intensity infection: 1 – 49 *S. haematobium* eggs/ 10 ml urine [43], based on the arithmetic mean egg counts of all samples provided by a study participant pre- or post-MDA. Heavy intensity infection: ≥50 *S. haematobium* eggs/ 10 ml urine [43], based on the arithmetic mean egg counts of all samples provided by a study participant pre- or post-MDA.

Fig 3 shows the change in infection intensity classes from pre- to post-MDA in each school. In School 1, all students (100%; 50/50) that had a light intensity *S. haematobium* infection pre-MDA were negative post-MDA. Most students (83.3%; 10/12) that had a heavy intensity infection pre-MDA were also negative post-treatment. However, a couple (16.7%; 2/12) were found to have a light intensity infection post-MDA. In School 2, most students (90.2%; 148/164) who had a light intensity infection pre-MDA were negative post-treatment, but several (9.8%; 16/164) remained lightly infected. Among the heavily infected students pre-MDA in School 2, less than half (40.6%; 28/69) were found to be negative post-treatment, with all the remaining students either presenting with a light (53.6%; 37/69) or a heavy (5.8%; 4/69) intensity infection.

**Fig 3.**
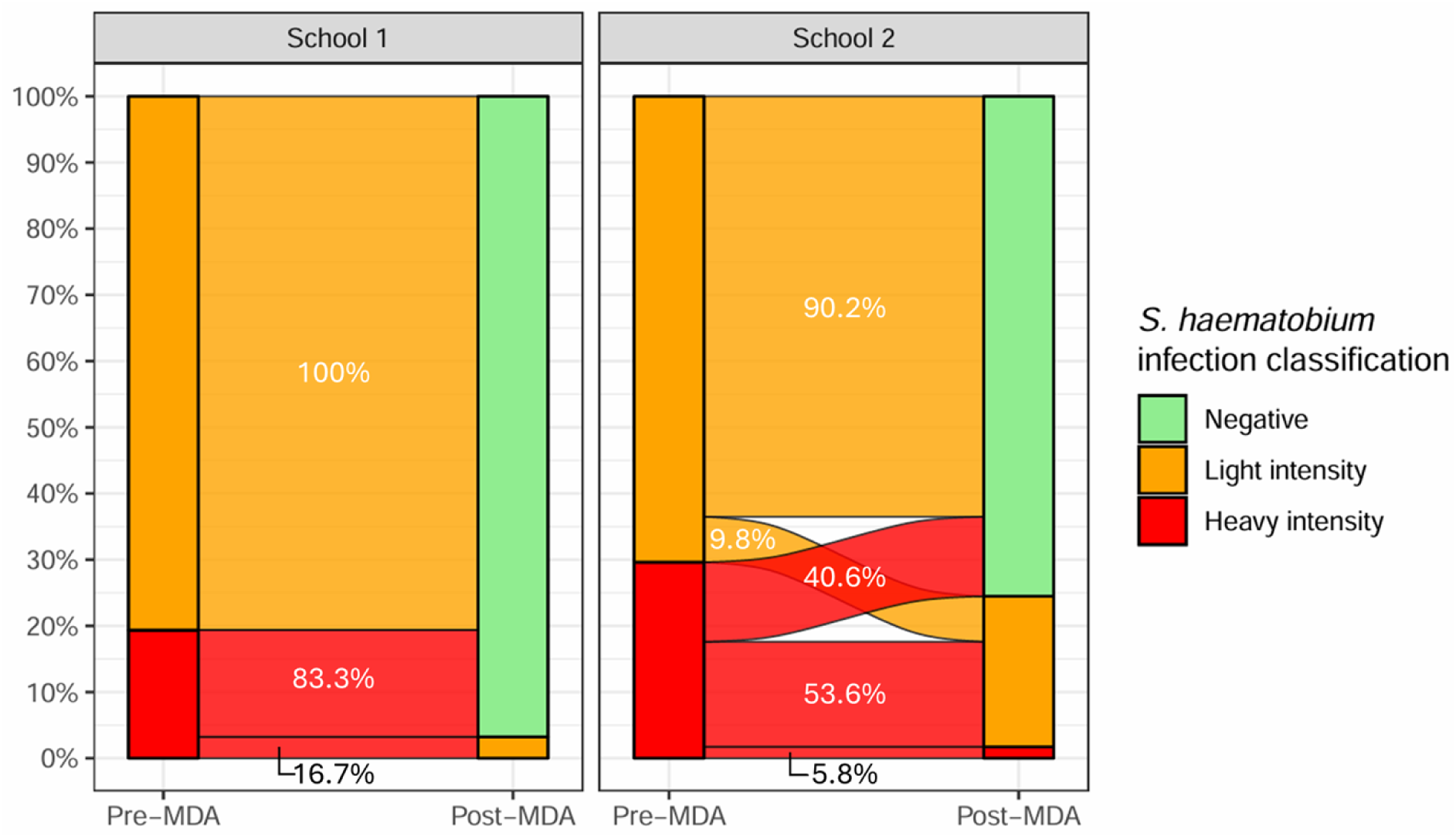
Change in *Schistosoma haematobium* infection intensity pre- and post-MDA for students with confirmed infection pre-MDA. The percentages in the flows represent the proportions of post-MDA classification categories per pre-MDA classification category. Light intensity infection: 1 – 49 *S. haematobium* eggs/ 10 ml urine [43], based on the arithmetic mean egg counts of all samples provided by a study participant pre- or post-MDA. Heavy intensity infection: ≥50 *S. haematobium* eggs/ 10 ml urine [43], based on the arithmetic mean egg counts of all samples provided by a study participant pre- or post-MDA.

### Drug efficacy of praziquantel

The primary outcome measure for drug efficacy, ERRs based on arithmetic mean eggs counts, were 99.8% (90% CI: 99.2-100%) in School 1 and 94.8% (90% CI: 88.6-98.6%) in School 2. The test for adequate drug efficacy showed that efficacy was significantly non-inferior to 93% for School 1, but not for School 2 (Fig 4A). The test for reduced drug efficacy failed to demonstrate that efficacy was significantly inferior to 98% for both schools. Hence, drug efficacy in terms of ERRs was considered “adequate” for School 1 but “inconclusive” for School 2.

**Fig 4.**
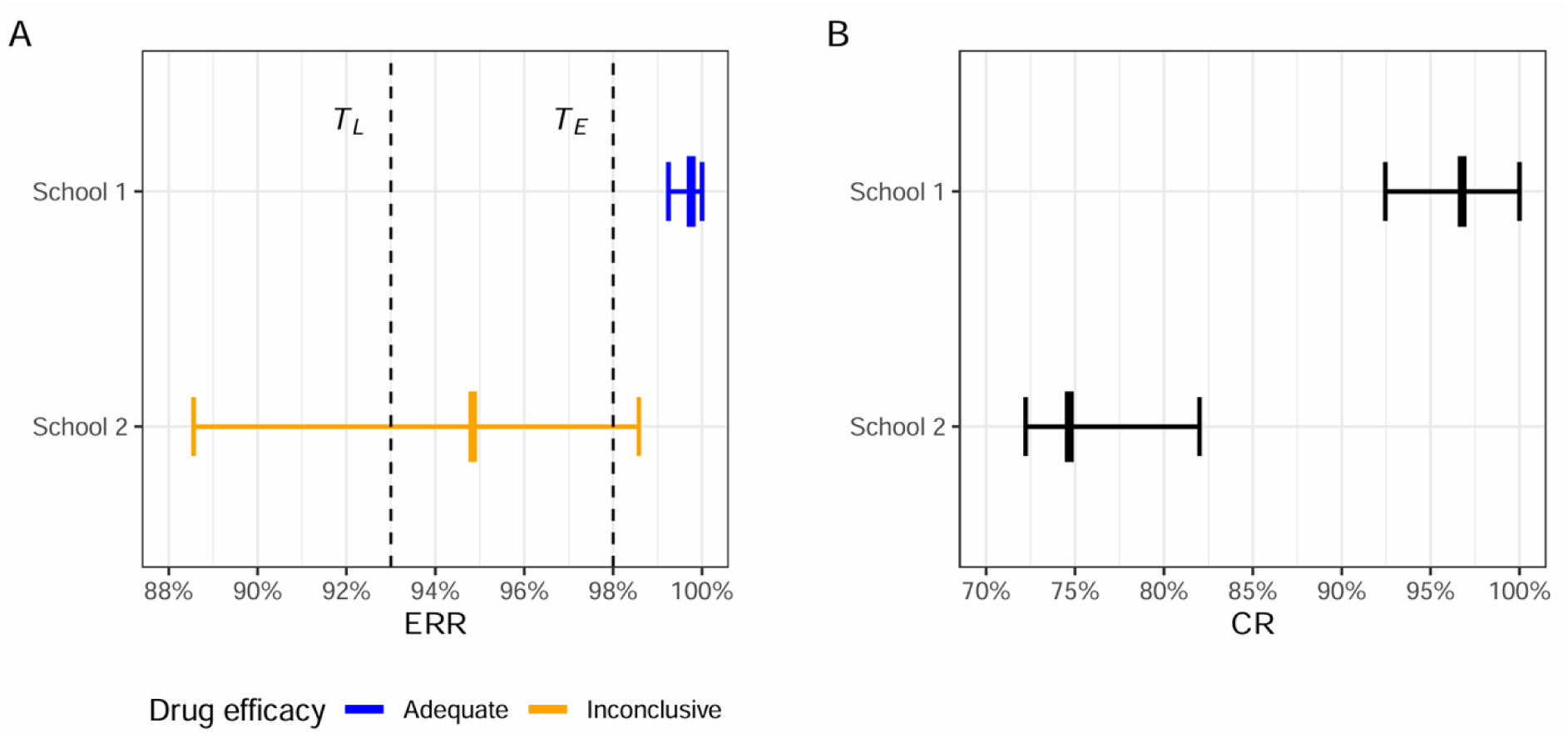
Point estimates and confidence intervals of egg reduction rates and cure rates after praziquantel treatment in two schools in Pemba, Tanzania. The point estimates of the egg reduction rates (ERRs) (A) and cure rates (CRs) (B) are indicated by the bold vertical markers, with the horizontal bars representing the 90% CIs calculated by bootstrapping. For School 1, the ERR was not significantly inferior (upper confidence limit was above *T_E_*) and the non-inferiority test was statistically significant (lower confidence limit was above *T_L_*), indicating adequate drug efficacy. For School 2, the null hypotheses of both the inferiority test and non-inferiority test could not be rejected (upper confidence limit was above *T_E_* and lower limit was below *T_L_*, respectively), meaning the overall result was inconclusive. *T_E_*: expected drug efficacy; *T_L_*: lower drug efficacy threshold.

In addition to ERR, we also determined the CRs (Fig 4B). In School 1, the CR was 96.8% (90% CI: 92.4 – 100%), and in School 2, the CR was 74.7% (90% CI: 72.2 – 82.0%).

### Association between individual pre-MDA egg intensity and egg reduction rate

The median posterior ERR as estimated by the GLMM for the population treatment effect was 100% (95% BCI, 99.8 – 100%) for School 1 and 99.8% (95% BCI, 99.6 – 99.8%) for School 2 (Fig 5, dashed grey lines and bands). These ERRs are conditional on the lognormal-distributed random effects in the model and therefore represent a reduction in the geometric mean count (as opposed to the data-derived ERRs that represent a reduction in the arithmetic mean count). The GLMM with individual-specific treatment effects generated a posterior for each individual ERR in geometric mean count. The medians of these individual ERR posteriors (dots in Fig 5) had a median value of 100% (interquartile range, 100 – 100%) in School 1 and 100% (interquartile range, 99.9 – 100%) in School 2. For School 1, the model estimated a posterior mean *β*_1_ of -0.004 (95% BCI, -0.083 – 0.066). As the *β*_1_ posterior mass covered values both below and above zero, an association between treatment efficacy and pre-MDA infection intensity was unlikely for School 1 (Fig 5, horizontal blue regression line for School 1). In School 2, the posterior mean *β*_1_ was -0.048 (95% BCI, -0.091 – -0.013). With 99.7% of the posterior mass of *β*_1_ < 0, there was a high likelihood of a negative association between pre-MDA infection intensity and treatment effect in School 2. Consequently, compared to the median individual ERR of 100% in School 2, the individual with the highest estimated pre-MDA egg count in School 2 had a lowered predicted ERR of 99.4%.

**Fig 5.**
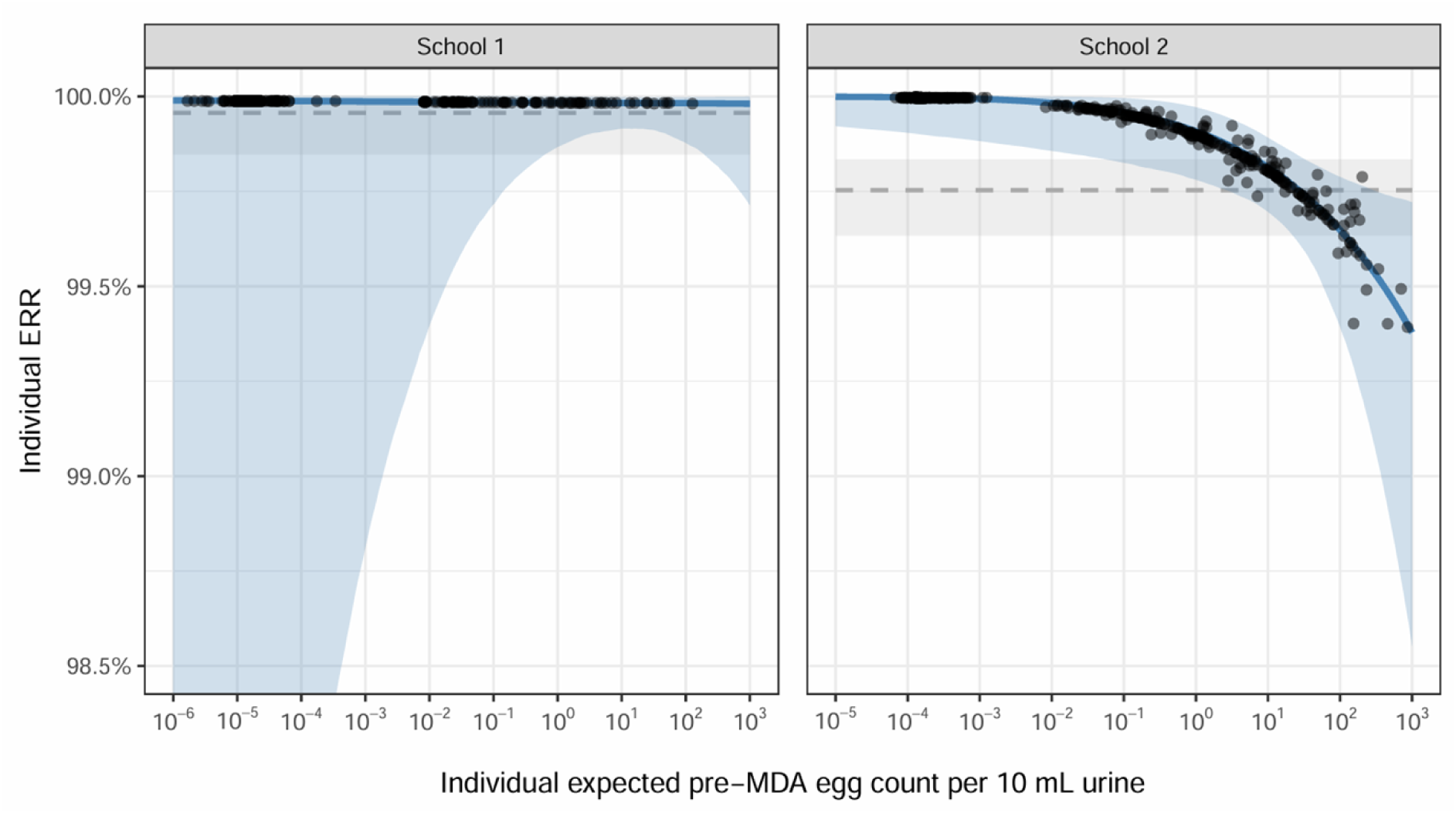
Individual egg reduction rates as a function of the expected pre-MDA egg count in two schools in Pemba, Tanzania. The posterior mean regression lines (blue line) with 95% credible bands (blue shaded area) were generated with the generalized linear mixed model with individual treatment effect as a function of the individual random effect, which was converted here to the latent individual pre-MDA egg count per 10 mL urine. The dots mark the model-estimated pre-MDA egg counts of the school students (based on their individual random effect and mean of day random effects, with the latter causing the vertical jitter from the regression line) *versus* their individual egg reduction rate (ERR). The dashed lines and grey bands indicate the ERRs and 95% credible bands, respectively, estimated by the generalized linear mixed model with the common population treatment effect.

## Discussion

As part of the RESIST project [35], we assessed the drug efficacy of praziquantel (40 mg/kg), used on the Zanzibar islands for regular MDA for more than 20 years, in two hotspot schools on Pemba. Using a sampling approach examining up to six urine samples per person pre- and post-MDA, we found a very high ERR of 99.8% in School 1 and a considerably lower ERR of 94.8% in School 2. Both results are consistent with those of a recent meta-analysis, which indicated that with a praziquantel treatment dose of 40 mg/kg, an ERR of 95% can be achieved, regardless of the *Schistosoma* species treated [7]. Our results are also in line with other systematic reviews and meta-analyses that indicated ERRs of 92-98% [44–46]. According to the current WHO manual for drug efficacy monitoring [13], our finding of ERRs ≥90% (based on point-estimates) suggests that praziquantel (40 mg/kg) is still efficacious after more than 20 years of regular application in MDAs on the Zanzibar islands. However, importantly, the WHO-recommended approach is based on point-estimates [13], which is statistically invalid. Therefore, we extended our evaluations by formal hypothesis testing as per Denwood *et al*. [41], using a framework that is currently also used to update the WHO manual for drug efficacy monitoring. This hypothesis testing indicated that drug efficacy was “adequate” in School 1 but “inconclusive” in School 2, due to a too high uncertainty in the estimated ERR for School 2. Hence, in a strict statistical sense, reduced efficacy cannot be ruled out in School 2. Further research is required to investigate resistance as a potential cause of these findings. In the larger frame of the RESIST project, whole genome sequencing analyses of miracidia that were hatched from *S. haematobium* egg-positive urine samples post-MDA in both schools, and a specific focus on detecting potential mutations in the gene encoding the praziquantel drug target in *S. haematobium* (*Sh*TRPM_PZQ_ channel), will shed more light on genetic signatures that may point to reduced praziquantel efficacy in *S. haematobium* on Pemba [35].

While it is important to rule out the potential development of drug resistance at molecular level, a more evident explanation for the inconclusive ERR observed here may be the higher pre-MDA infection intensities in School 2. Before MDA, the proportion of heavy intensity *S. haematobium* infections was considerably higher in School 2 than in School 1 (11.2% *versus* 2.8%). Individuals that were still egg-positive post-MDA mostly originated from the high intensity groups pre-MDA. The GLMM analyses confirmed that higher pre-MDA infection intensity was associated with a moderate reduction in individual ERR in School 2. This finding contrasts with a previous meta-analysis of praziquantel efficacy for *S. mansoni* and *S. haematobium* (combined in one analysis), which found that ERR increased, rather than decreased, with increasing pre-treatment infection intensity [7]. Another study used data from nine schistosomiasis intervention studies in Africa in a conditional mixed model and found no statistically significant relationship between an individual’s egg count before treatment and their ERR [47]. There are several hypotheses that could explain how heavy intensity infections in our study were associated with lower ERR. First, heavily infected individuals may be strongly exposed to infection and hence harbor many juvenile worms, which are refractory to praziquantel and will mature to reproducing adults after treatment. Second, a single dose of praziquantel may only be able to kill up to finite number of adult worms, meaning that the fraction of worms killed might be lower in heavily infected individuals. Lastly, individuals with heavy infections may have an initially larger number of eggs trapped and passing through the tissues, many of which may continue to be released for some time after treatment.

In addition to the primary endpoint of ERRs, we also evaluated CRs for the sake of comparison to existing literature. We found CRs of 96.8% in School 1 and of 74.7% in School 2. Like the ERRs, these results were in line with a recent meta-analysis which reported a synthesized CR of 57-88%, depending on schistosome species and number of samples examined [7]. Similarly, several other meta-analyses report CRs of 64-81% for urogenital schistosomiasis [44–46].

Notably, the CRs in the current study were considerably higher than results from a drug efficacy study conducted on Pemba in 2007, where the CR was only 46% three weeks post-treatment [48]. Importantly, we would like to highlight that CR is considered of limited use to monitor drug efficacy in population surveys since it is influenced by i) the intensity of infection at baseline and ii) the sensitivity of the method used to diagnose the infection [14]. Additionally, we would like to stress that CRs are mostly used in clinical trials, which differ from drug efficacy monitoring studies in two important ways: i) clinical trials often select only egg-positive individuals for follow-up, leading to an overestimation of drug efficacy (ERR and CR) due to regression towards the mean, which drug efficacy monitoring studies should avoid [49,50]; and ii) clinical trials often employ intention-to-treat analyses, which can lead to underestimation of drug efficacy due to non-compliance of study participants. Hence, while our stringent sampling approach will have reduced the probability of overestimating the CR in our study, the direct comparison of CRs identified in our research with the results of other studies that used different study designs and methodological approaches needs to be interpreted with care.

Our study presents several strengths but also certain limitations. The repeated urine sampling approach was more sensitive for detection of egg-excreting individuals compared to standard single-day sampling [51,52]. It also enabled correction for day-to-day variation in our GLMM-based ERR estimation. It should be noted that for School 1, the majority of students in the PP cohort submitted fewer than five urine samples post-MDA. In itself, we reason that this should have only affected the statistical accuracy of our analysis; we do not expect that it has biased our estimates of the ERRs, as it seems unlikely that failure to deliver a urine sample post-MDA on a given day is correlated with higher egg count in the urine on that same day. Ongoing modelling analyses as part of the RESIST project will further elucidate the influence of sampling effort on the accuracy of drug efficacy monitoring.

We conclude that after more than 20 years of regular MDA, the efficacy of praziquantel (40 mg/kg) on the Zanzibar islands remains high, even higher than the flawed 90% ERR threshold for optimal efficacy suggested by the WHO [13]. Although the inconclusive efficacy results for School 2 are most likely caused by high intensity infections pre-MDA that were associated with a lower individual ERRs, more in-depth investigation is needed. Future molecular work of the RESIST project will show whether the *S. haematobium* miracidia isolated from egg-positive individuals post-MDA carry genetic markers of reduced praziquantel susceptibility. Ongoing genome wide analysis of the *S. haematobium* miracidia collected from the same study participants pre- and post-MDA, as part of the overall RESIST project, aims to characterise praziquantel susceptibility of these *S. haematobium* populations at the molecular level by genotyping variants and functionally profiling activity of the target *Sh*TRPM_PZQ_ [20,25], deciphering between signs of praziquantel resistance at the genetic level versus other treatment deficiencies.

## Data Availability

The minimal data set and accompanying dictionary is available as supporting information of this manuscript and will be published on at the Infectious Disease Data Observatory (IDDO) https://www.iddo.org/.

## Acknowledgements

We acknowledge the great support of the Zanzibar Ministry of Health and of community leaders, principals, teachers and parents on Pemba for this study. We are indebted to the students who participated in our study and submitted multiple urine samples before and after mass drug administration (MDA). We are grateful for the dedicated and high-quality work of the field and laboratory research teams from the Public Health Laboratory-Ivo de Carneri. We thank the members of the Pemba Neglected Tropical Diseases Program from the Zanzibar Ministry of Health for aligning the MDA in the two study schools with the timeline of the RESIST project and for their great support during the MDA. Moreover, we thank Unlimit Health for supporting the MDA implementation. Last but not least, we are grateful to the advisory board of the RESIST project for their interest and support of the study.

## Author Contributions

Stefanie Knopp: Conceptualization, Data Curation, Funding Acquisition, Investigation, Methodology, Project Administration, Resources, Supervision, Writing – Original Draft Preparation, Writing – Review & Editing

Norbert J. van Dijk: Data Curation, Formal Analysis, Writing – review & Editing

Naomi C. Ndum: Investigation, Writing – Review & Editing

Tom Pennance: Investigation, Writing – Review & Editing

Mohammed N. Ali: Investigation

Khamis R. Suleiman: Investigation

Matthew Denwood: Methodology, Writing – Review & Editing

Saleh Juma: Resources, Supervision

Shaali M. Ame: Resources

Aidan M. Emery: Conceptualization, Funding Acquisition, Writing – Review & Editing

Bonnie Webster: Conceptualization, Funding Acquisition, Investigation, Writing – Review & Editing

Luc E. Coffeng: Conceptualization, Funding Acquisition, Methodology, Project Administration, Resources, Writing – Review & Editing

Said Mohammed Ali: Conceptualization, Funding Acquisition, Investigation, Project Administration, Resources, Supervision

## Supporting information

**S1 STROBE Checklist.**

**(PDF)**

**S1 Minimal dataset and dictionary.**

**(XLSX)**

**S1 Fig. Anthelmintic drug efficacy classification framework.**

**(PDF)**

